# Household overcrowding and risk of SARS-CoV-2: analysis of the Virus Watch prospective community cohort study in England and Wales

**DOI:** 10.1101/2021.05.10.21256912

**Authors:** Robert W Aldridge, Helen Pineo, Ellen Fragaszy, Max Eyre, Jana Kovar, Vincent Nguyen, Sarah Beale, Thomas Byrne, Anna Aryee, Colette Smith, Delanjathan Devakumar, Jonathon Taylor, Srinivasa Vittal Katikireddi, Wing Lam Erica Fong, Cyril Geismar, Parth Patel, Madhumita Shrotri, Isobel Braithwaite, Annalan M D Navaratnam, Anne M Johnson, Andrew Hayward, on behalf of Virus Watch Collaborative

**Affiliations:** Centre for Public Health Data Science, Institute of Health Informatics, University College London, UK; Institute for Environmental Design and Engineering, Bartlett School of Environment, Energy and Resources, University College London, London, UK; Institute of Epidemiology and Health Care, University College London, London, UK; Department of Infectious Disease Epidemiology, London School of Hygiene and Tropical Medicine, Keppel Street, London, UK; Centre of Health Informatics, Computing and Statistics, Lancaster Medical School, Lancaster University, Lancaster, LA1 4YW, UK; Liverpool School of Tropical Medicine, Liverpool, UK; Institute for Global Health, University College London, London, UK; Department of Civil Engineering, Tampere University, Finland; MRC/CSO Social and Public Health Sciences Unit, University of Glasgow Institute of Health and Wellbeing, Glasgow, UK; Department of Population, Policy and Practice, UCL Great Ormond Street Institute of Child Health, London, UK; Francis Crick Institute, London, UK; Health Protection and Influenza Research Group, Division of Epidemiology and Public Health, University of Nottingham School of Medicine, Nottingham, United Kingdom; University College London Hospital, London, United Kingdom; Department of Computer Science, University College London, London, UK; Centre for Behaviour Change, University College London, London, UK; London Centre for Nanotechnology and Division of Medicine, London, UCL; SpaceTimeLab, Department of Civil, Environmental and Geomatic Engineering, University College London, London, UK; Royal Free London NHS Foundation Trust, London, UK

## Abstract

**Background:** Household overcrowding is associated with increased risk of infectious diseases across contexts and countries. Limited data exist linking household overcrowding and risk of COVID-19. We used data collected from the Virus Watch cohort to examine the association between overcrowded households and SARS-CoV-2.

**Methods:** The Virus Watch study is a household community cohort of acute respiratory infections in England & Wales that began recruitment in June 2020. We calculated the persons per room for each household and classified accommodation as overcrowded when the number of rooms□was fewer than the number of people. We considered two primary outcomes - PCR-confirmed positive SARS-CoV-2 antigen tests and laboratory confirmed SARS-CoV-2 antibodies (Roche Elecsys anti-N total immunoglobulin assay). We used mixed effects logistic regression models that accounted for household structure to estimate the association between household overcrowding and SARS-CoV-2 infection.

**Results:** The proportion of participants with a positive SARS-CoV-2 PCR result was highest in the overcrowded group (6.6%; 73/1,102) and lowest in the under-occupied group (2.9%; 682/23,219). In a mixed effects logistic regression model that included age, sex, ethnicity, household income and geographical region, we found strong evidence of an increased odds of having a positive PCR SARS-CoV-2 antigen result (Odds Ratio 3.72; 95% CI: 1.92, 7.13; p-value < 0.001) and increased odds of having a positive SARS-CoV-2 antibody result in individuals living in overcrowded houses (2.96; 95% CI: 1.13, 7.74; p-value =0.027) compared to people living in under-occupied houses. The proportion of variation at the household level was 75.1% and 74.0% in the PCR and antibody models respectively.

**Discussion:** Public health interventions to prevent and stop the spread of SARS-CoV-2 should consider the much greater risk of infection for people living in overcrowded households and pay greater attention to reducing household transmission. There is an urgent need to better recognise housing as a leading determinant of health in the context of a pandemic and beyond.

## Introduction

Household overcrowding is associated with increased risk of infectious diseases across cultures and countries.^1^ The World Health Organization Housing and Health Guidelines emphasise the health risks of overcrowding and note its complex economic, social and political determinants.^2,3^ Several definitions of overcrowding exist and it is also used as an indicator of material deprivation.^4^ Measures of overcrowding assess whether there is adequate dwelling space for occupants’ needs related to shelter, space and privacy.^1^

According to the English Housing Survey, approximately 787,000 (3%) of English households are overcrowded with unequal distribution across regions and social groups.^5^ Seven percent of the most deprived households were overcrowded compared with less than half a percent of the least deprived households. Overcrowding was highest in London compared to all other English regions. White British households are less likely to be overcrowded than households from all other ethnic groups.

There is strong evidence that household overcrowding is associated with risk of infectious diseases such as tuberculosis,^1^ and increasing evidence on the association between household overcrowding and COVID-19. In the USA, an ecological analysis found that COVID-19 death rates were higher in the counties with highest percentage of household crowding (16.8 per 100,000) compared to the least crowded areas (4.9 per 100,000).^6^ A study using 2011 UK census data examined ethnicity, household composition and COVID-19 mortality and found that elderly adults living with younger people were at increased risk of COVID-19 mortality.^7^ The study described the distribution of overcrowded housing within census data used, but it did not adjust for overcrowded housing in the causal mediation analysis as these were considered consequences of living in a multi-generational household rather than confounding factors. A study of UK Biobank data found household size was associated with COVID-19 infection after adjusting for age, sex, deprivation, ethnicity and Body Mass Index (BMI).^8^ The REACT study found SARS-CoV-2 antibodies increased from 4.7% for people in single person households to 13% in households of seven or more.^9,10^ The OpenSAFELY study found household size accounted for 10-16% of the excess risk of testing positive for SARS-CoV-2 and 12-39% of the excess risk of COVID-19 mortality in South Asian groups.^11^

Relatively few studies have investigated the impact of overcrowding specifically, as opposed to household size. In Birmingham (UK), an analysis of data from 408 hospitalised COVID-19 patients found that people from areas of the city with low housing quality and overcrowding were more likely to be admitted to intensive care compared to other areas.^12^ A study of ≥70 year olds using Swedish cause of death, administrative and dwelling registry data corroborates this finding with individual-level overcrowding data. After adjusting for individual age, sex, country of birth, income and education, COVID-19 mortality was 2.1 times (HR 2.1, 95% CI 1.53-2.87) higher among ≥70 year olds in households with less than 20m^2^/inhabitant compaired with those with >60m^2^/inhabitant.^13^ In a serological survey undertaken in Lima, Peru, lateral-flow SARS-CoV-2 seropositivity was also more prevalent among households in the two most overcrowded quartiles compared with the least overcrowded quartile (respective prevalence ratios 1.41 & 1.99, 95%CI 1.01-1.97 and 1.41-2.81) as measured by a ratio of inhabitants to habitable rooms with adjustment for sex, age group, local region and socio-economic status.^14^ Overcrowding as defined by more than 3 household members with fewer than six rooms between them carried increased odds of laboratory-confirmed secondary infection of SARS-CoV-2 in 92 North Carolinan households within 28 days of a PCR-confirmed index case. Higher rates of household overcrowding observed among non-White/Hispanic were also argued to account for the higher rates of secondary tranmission found among those ethnic groups in this study.^15^

Additional evidence on the association between household overcrowding and COVID-19 risk can guide immediate public health mitigation measures to reduce the spread of the SARS-CoV-2 and inform long-term housing policy in the UK. We used data collected from the Virus Watch cohort to examine the association between overcrowded households and either Polymerase Chain Reaction (PCR) confirmed SARS-CoV-2 or antibodies to acquired through SARS-CoV-2 infection.^16^

## Methods

The Virus Watch study is a household community cohort of acute respiratory infections in England & Wales that started recruitment in June 2020.^16^ As of 3rd May 2021, there were 50,648 participants in Virus Watch that were recruited using a range of methods including post, social media, and SMS messages and letters from their General Practices (Table 1). Participants were followed-up weekly by email with a link to an illness survey which asks about the presence or absence of symptoms that could indicate COVID-19 disease including respiratory, gastrointestinal and general infection symptoms. The weekly survey is also used to capture SARS-CoV-2 test results received from outside the study (eg. via the UK Test-Trace-Isolate system).

**Table 1.**
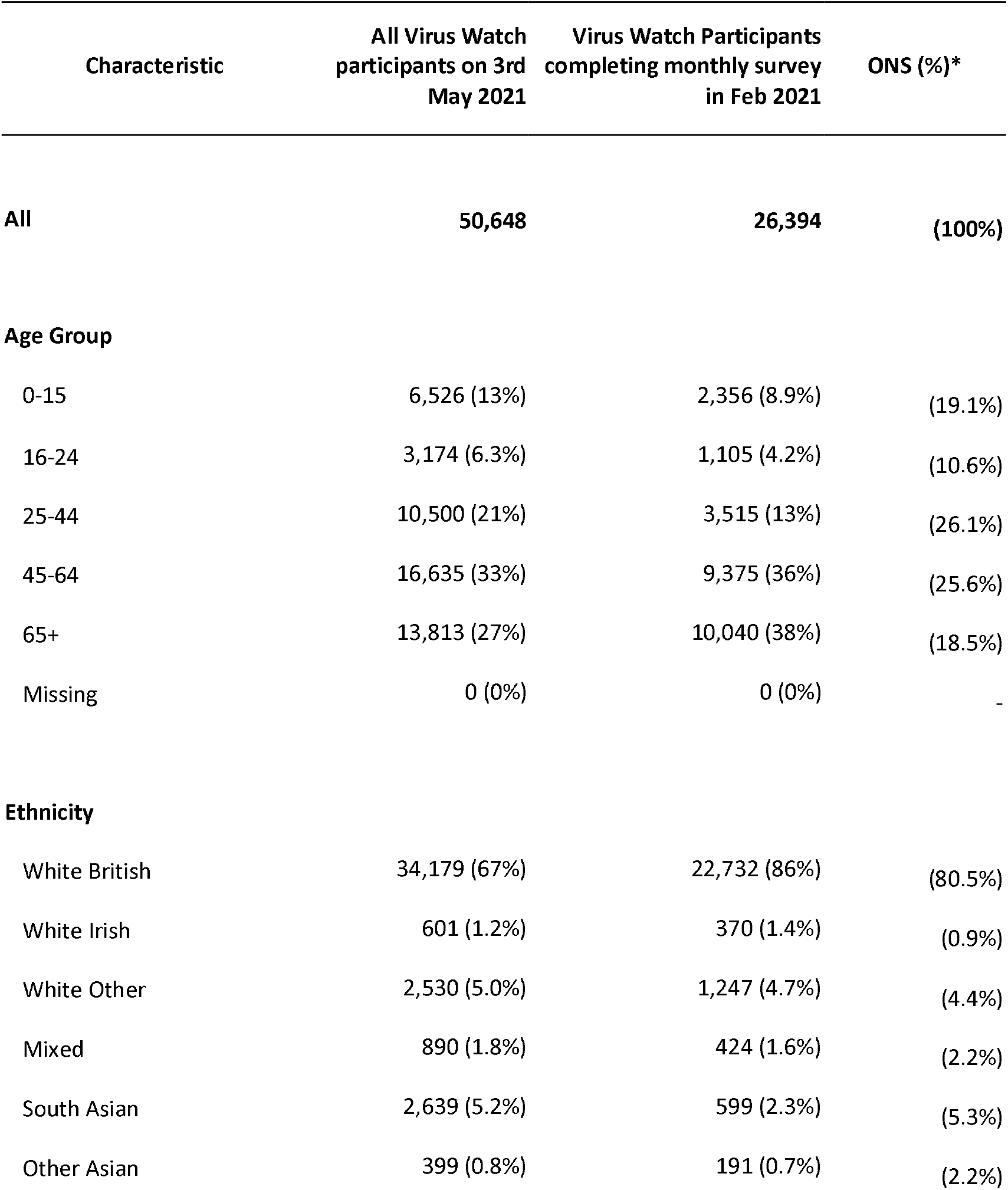
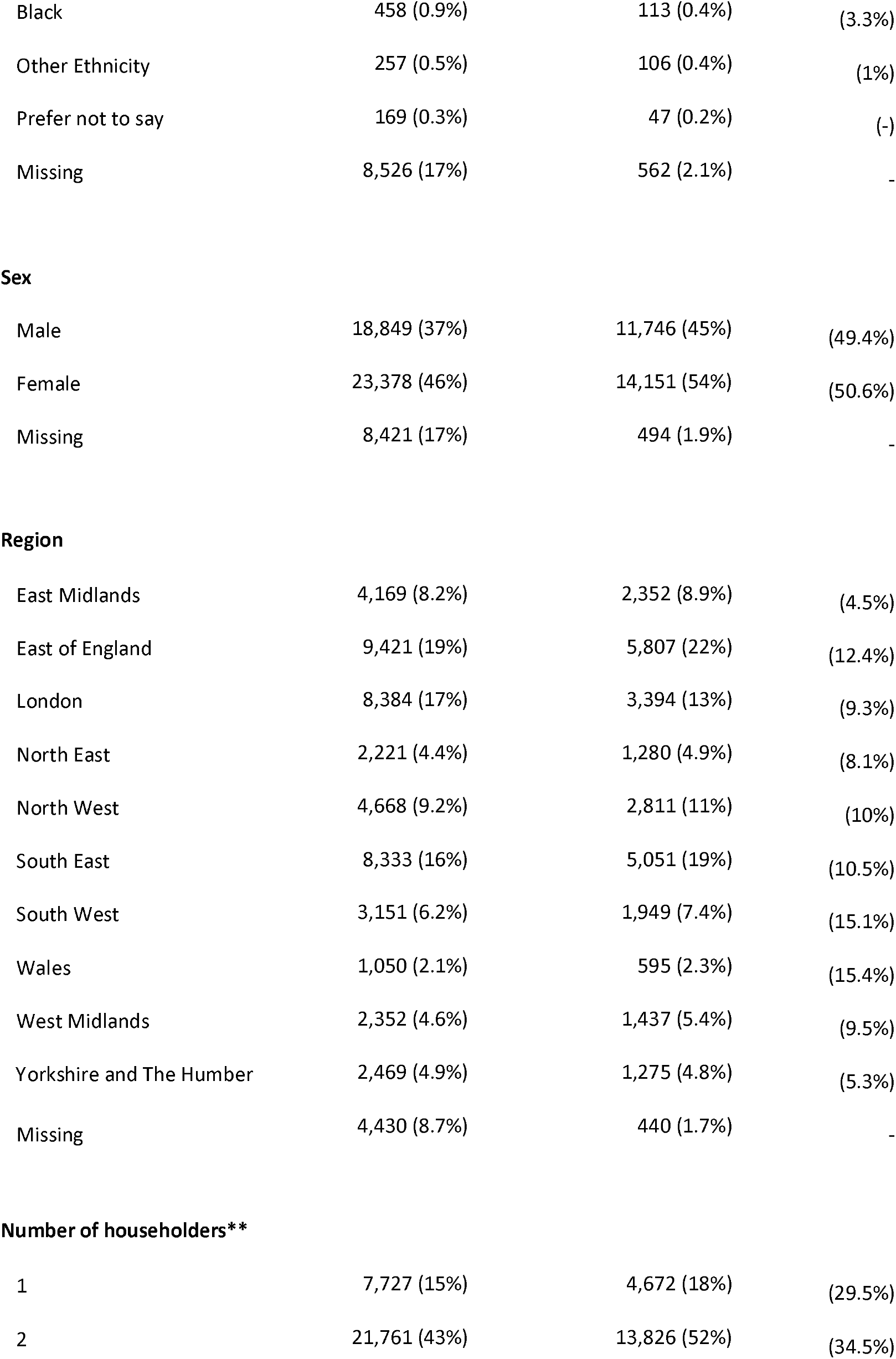
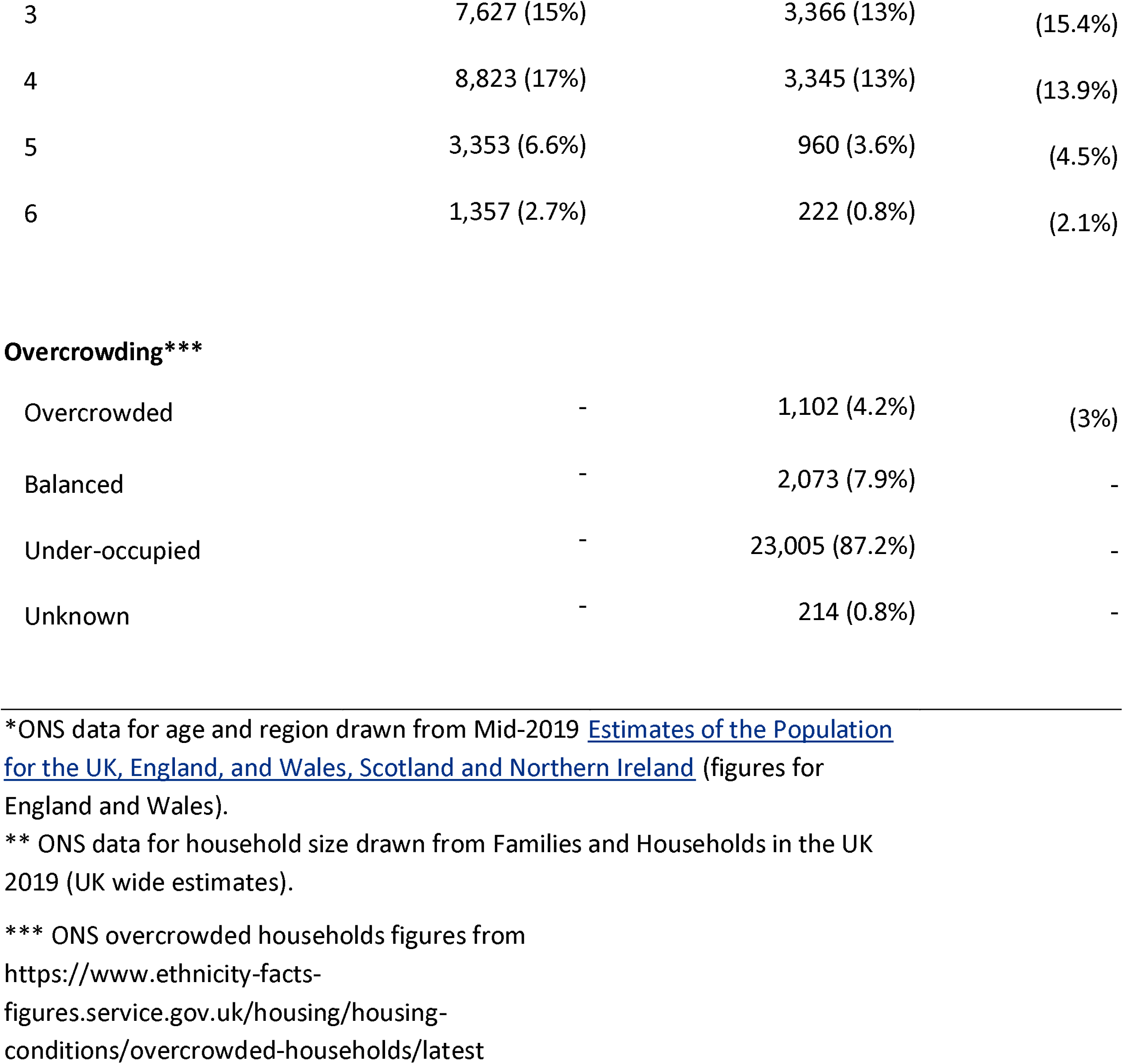
Description of Virus Watch participants who took part in the February 2021 survey compared to the whole Virus Watch cohort on 3rd May 2021 and ONS census data for England and Wales.

| Characteristic | All Virus Watch participants on 3rd May 2021 | Virus Watch Participants completing monthly survey in Feb 2021 | ONS (%)* |
| --- | --- | --- | --- |
| **All** | **50,648** | **26,394** | **(100%)** |
| **Age Group** |  |  |  |
| 0-15 | 6,526 (13%) | 2,356 (8.9%) | (19.1%) |
| 16-24 | 3,174 (6.3%) | 1,105 (4.2%) | (10.6%) |
| 25-44 | 10,500 (21%) | 3,515 (13%) | (26.1%) |
| 45-64 | 16,635 (33%) | 9,375 (36%) | (25.6%) |
| 65+ | 13,813 (27%) | 10,040 (38%) | (18.5%) |
| Missing | 0 (0%) | 0 (0%) | - |
| **Ethnicity** |  |  |  |
| White British | 34,179 (67%) | 22,732 (86%) | (80.5%) |
| White Irish | 601 (1.2%) | 370 (1.4%) | (0.9%) |
| White Other | 2,530 (5.0%) | 1,247 (4.7%) | (4.4%) |
| Mixed | 890 (1.8%) | 424 (1.6%) | (2.2%) |
| South Asian | 2,639 (5.2%) | 599 (2.3%) | (5.3%) |
| Other Asian | 399 (0.8%) | 191 (0.7%) | (2.2%) |
| Black | 458 (0.9%) | 113 (0.4%) | (3.3%) |
| Other Ethnicity | 257 (0.5%) | 106 (0.4%) | (1%) |
| Prefer not to say | 169 (0.3%) | 47 (0.2%) | (-) |
| Missing | 8,526 (17%) | 562 (2.1%) | - |
| Sex |  |  |  |
| Male | 18,849 (37%) | 11,746 (45%) | (49.4%) |
| Female | 23,378 (46%) | 14,151 (54%) | (50.6%) |
| Missing | 8,421 (17%) | 494 (1.9%) | - |
| Region |  |  |  |
| East Midlands | 4,169 (8.2%) | 2,352 (8.9%) | (4.5%) |
| East of England | 9,421 (19%) | 5,807 (22%) | (12.4%) |
| London | 8,384 (17%) | 3,394 (13%) | (9.3%) |
| North East | 2,221 (4.4%) | 1,280 (4.9%) | (8.1%) |
| North West | 4,668 (9.2%) | 2,811 (11%) | (10%) |
| South East | 8,333 (16%) | 5,051 (19%) | (10.5%) |
| South West | 3,151 (6.2%) | 1,949 (7.4%) | (15.1%) |
| Wales | 1,050 (2.1%) | 595 (2.3%) | (15.4%) |
| West Midlands | 2,352 (4.6%) | 1,437 (5.4%) | (9.5%) |
| Yorkshire and The Humber | 2,469 (4.9%) | 1,275 (4.8%) | (5.3%) |
| Missing | 4,430 (8.7%) | 440 (1.7%) | - |
| Number of householders\*\* |  |  |  |
| 1 | 7,727 (15%) | 4,672 (18%) | (29.5%) |
| 2 | 21,761 (43%) | 13,826 (52%) | (34.5%) |
| 3 | 7,627 (15%) | 3,366 (13%) | (15.4%) |
| 4 | 8,823 (17%) | 3,345 (13%) | (13.9%) |
| 5 | 3,353 (6.6%) | 960 (3.6%) | (4.5%) |
| 6 | 1,357 (2.7%) | 222 (0.8%) | (2.1%) |
| **Overcrowding\*\*\*** |  |  |  |
| Overcrowded | - | 1,102 (4.2%) | (3%) |
| Balanced | - | 2,073 (7.9%) | - |
| Under-occupied | - | 23,005 (87.2%) | - |
| Unknown | - | 214 (0.8%) | - |

\*ONS data for age and region drawn from Mid-2019 Estimates of the Population for the UK, England, and Wales, Scotland and Northern Ireland (figures for England and Wales).
\*\* ONS data for household size drawn from Families and Households in the UK 2019 (UK wide estimates).
\*\*\* ONS overcrowded households figures from https://www.ethnicity-facts-figures.service.gov.uk/housing/housing-conditions/overcrowded-households/latest

### Laboratory cohort

Nested within this larger study is a sub-cohort of 10,330 adults (aged over 18) participating in monthly antibody testing who completed at-home capillary blood sampling kits sent via post on a monthly basis, and provided self-reported vaccination data on a weekly basis, in addition to demographic and clinical data at baseline. Individuals were included in this analysis if they underwent antibody testing between 1st February 2021 and 3rd May 2021 and completed the February 2021 monthly survey.

### Monthly survey

The Virus Watch monthly survey includes demographic, psychosocial/behavioral, environmental and health-related questions beyond the scope of the weekly survey. Data used in this analysis are taken from the Virus Watch third monthly survey that was conducted between 09 February and 16 February 2021 and occurred during the third national lockdown for both England and Wales. In this survey we included a series of questions about participants’ housing status adapted from housing-related items in the 2011 England and Wales Census as this the most comprehensive study in terms of housing characteristics collected.^17^ These comprised of: accommodation type; whether accommodation was self-contained; number of rooms available for exclusive use by the household, excluding bathrooms, toilets, halls or landings, or rooms that can be used only for storage (e.g., cupboards); number of bedrooms (built or converted for use as bedrooms, even if not currently used as a bedroom); housing tenure and, if rented, details of the rental arrangement; and whether the accommodation had central heating. Additionally, we presented binary (yes/no) questions about whether participants’ accommodation had visible mould or fungus and/or damp spots on the walls or ceiling due to their association with respiratory tract infections.^18,19^ Housing-related survey questions are presented in full in the supplementary appendix.

### Overcrowding measure

To investigate overcrowding, we first calculated the persons per room^4^ for each household, which is defined as the number of household occupants divided by the number of rooms, excluding kitchen or bathrooms. We chose this measure because we wanted to examine the link between overcrowding and infectious risk as a result of the density of people in the house rather than the family and demographic (e.g. age and sex) relationships between them, such as the bedroom standard which often allows shared bedrooms for each pair of same-sex adolescents (10–20 years) and each pair of same or opposite sex children (under 10 years old).^4^ We did not ask participants to exclude kitchens from their reporting of the number of rooms in their accommodation and therefore we have assumed that all accommodation had a kitchen and subtracted one from the total number of rooms reported by each household. The persons per room variable was then categorised as: 1) under-occupied accommodation where the number of rooms was greater than the number of □people; 2) balanced accommodation where the number of rooms was equal to the number of people; 3) overcrowded accommodation where the number of roomslwas fewer than the number of people.

### Outcomes

We considered two primary outcomes. First, PCR-confirmed SARS-CoV-2; either self-reported or conducted as part of the Virus Watch laboratory cohort. This was assessed in an analysis including all Virus Watch participants who completed the February 2021 monthly survey. We assumed that participants not reporting either a positive or negative test result had never been a PCR-confirmed SARS-CoV-2 case. Second, we examined laboratory confirmed SARS-CoV-2 antibodies acquired through infection, among participants who underwent antibody testing as part of the VirusWatch study and who completed the February 2021 monthly survey. The main outcome variable was evidence of prior infection defined as a cut off index of 0.1 or more on the Roche Elecsys anti-N total immunoglobulin assay that measures seropositive for the Nucleocapsid protein.

### Covariates

Our analysis strategy was informed by conceptual models^20,21^ that have previously described the possible pathways between ethnicity and socio-economic status, overcrowding and risk of SARS-CoV-2 infection and we developed a Directed Acyclic Graph to inform covariate selection (Supplementary Appendix Figure S1). We used this and the rules outlined by VanderWeele as principles of confounder selection.^22^ In our primary analysis we considered age, sex, ethnicity, household income and geographical region as covariates (Model A). Age, sex, ethnicity and geographical region were derived from participants’ responses to demographic questions at study baseline. Household income was derived from the February 2021 monthly survey.

### Statistical Analyses

We undertook description of the characteristics of included participants. To model the association between the selected covariates and each outcome, we conducted univariable and multivariable analyses using mixed effects logistic regression models with a household-level random effect to account for household-level variation not explained by the covariates using the glmer function in R 4.0.3.

### Sensitivity Analyses

We have examined our assumption regarding kitchens in our calculation of overcrowding, by only excluding one room from households with 2 or more rooms in a sensitivity analysis. In addition to the primary mixed effects model, we ran three other separate sensitivity analyses (all accounting for household-level clustering): first a minimal sufficient adjustment set informed by our Directed Acyclic Graph (Model B); second a model with age, sex, households with children (a binary variable representing households with and without children), and number of close contacts outside the household as covariates (repeating an analysis previously reported prior to having occupational category or household income data available (Model C); third a model with age, sex, ethnicity, income and occupation as covariates (Model D).

### Ethics

The Virus Watch study has been approved by the Hampstead NHS Health Research Authority Ethics Committee. Ethics approval number - 20/HRA/2320.

### Role of the funding source

The study sponsor(s) had no role in study design; in the collection, analysis, and interpretation of data; in the writing of the report; and in the decision to submit the paper for publication. We confirm that all authors accept responsibility to submit for publication. For information security reasons, RWA, EF, ME, JC, VN, SB, TB, AA, WLEF, CG, PP, MS, AMDN and AH had full access to all individual level data in the study analyses and all other authors had access to aggregated data.

## Results

On 16th February 2021, participants in the Virus Watch study were invited to take part in the monthly survey and we were able to include responses to the survey for 26,394 participants in our primary analyses. The median number of rooms per household was 6 (interquartile range 5-7) and the median number of householders was 2 (interquartile range 1-3). 4.2% of participants (1,102/26,394) were classified as living in overcrowded households and 7.9% (2,073/26,394) in balanced households.

Since June 2020, 1,201 participants have had a positive SARS-CoV-2 antigen test either self-reported and tested through the national test and trace system or as part of the Virus Watch laboratory cohort. There were 867 participants with a PCR confirmed SARS-CoV-2 antigen test and data on their housing status (3.3%; 867/26,394). The proportion of participants with a positive SARS-CoV-2 antigen result was highest in the overcrowded group (6.6%; 73/1,102) and lowest in the under-occupied group (2.9%; 682/23,219; Table 2). In a mixed effects logistic regression model that included age, sex, ethnicity, household income and geographical region as fixed effects and a household-level random effect we found strong evidence of an increased odds of having a positive SARS-CoV-2 antigen result in individuals living in overcrowded accommodation (Odds Ratio (OR): 3.70; 95% CI: 1.92, 7.13; p-value < 0.001) and those in balanced homes (OR: 2.70; 95% CI: 1.68, 4.34; p-value < 0.001) compared to those in under-occupied homes (Table 3). The proportion of variation at the household level was 75.1%.

**Table 2.**
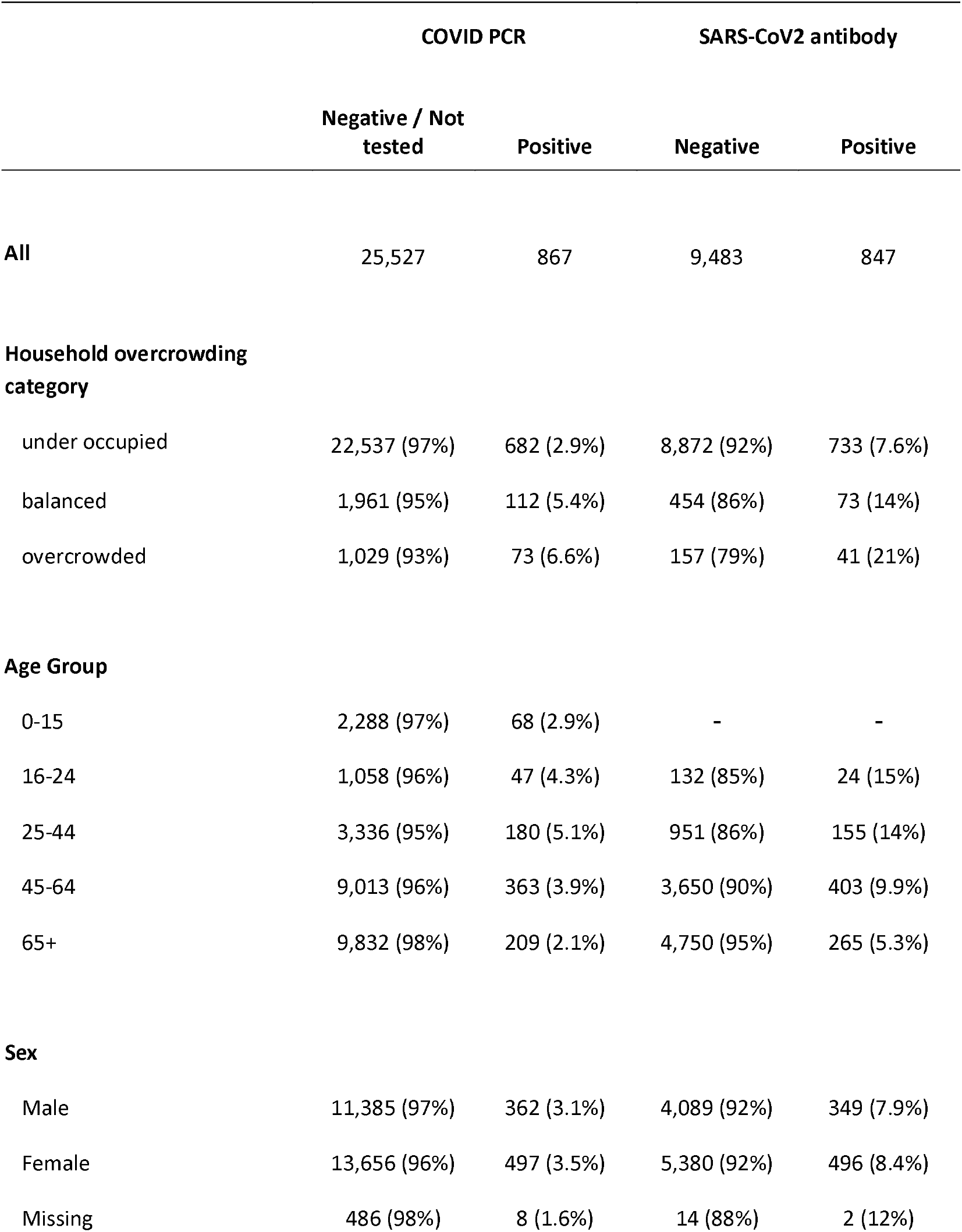
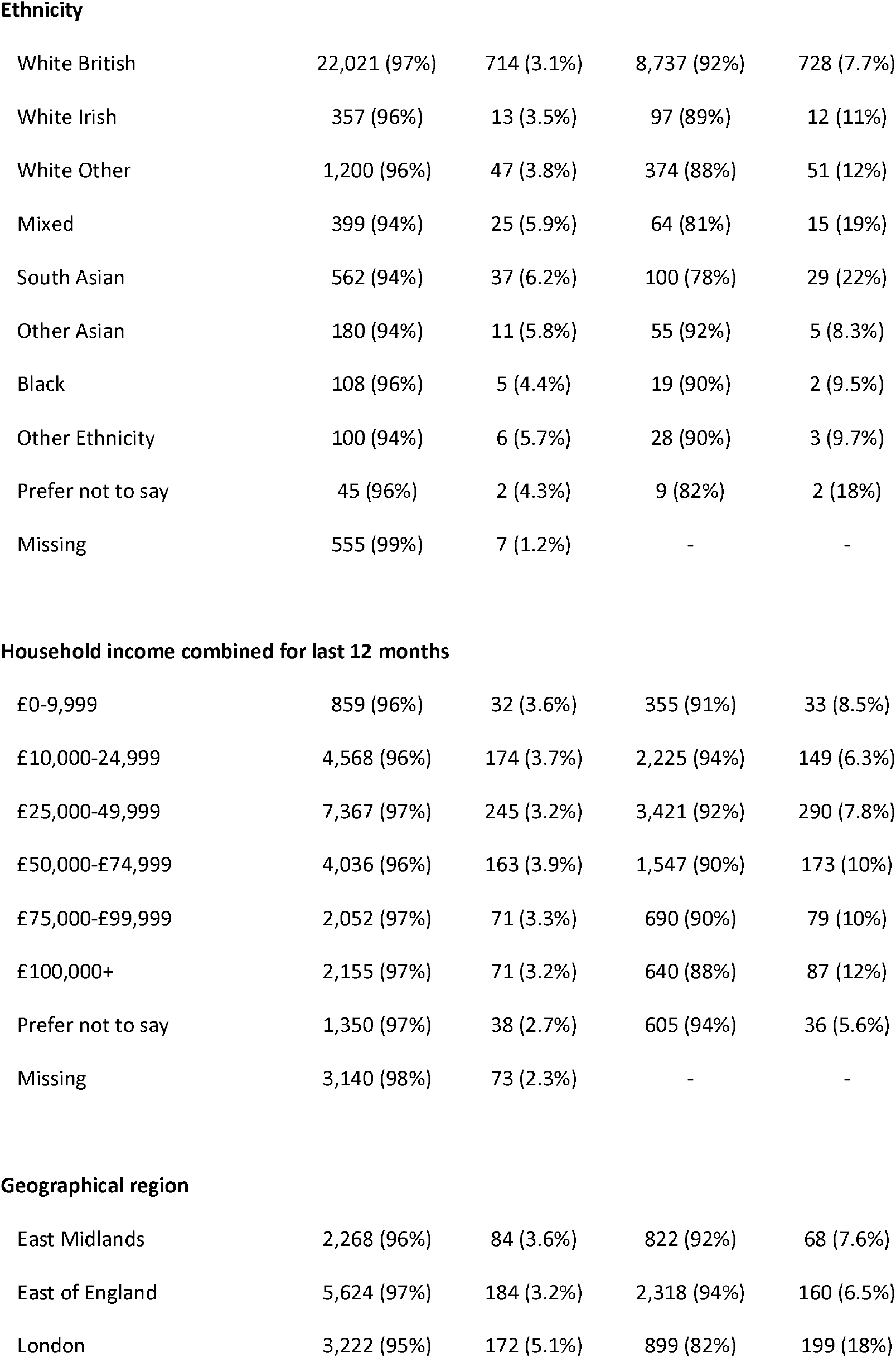
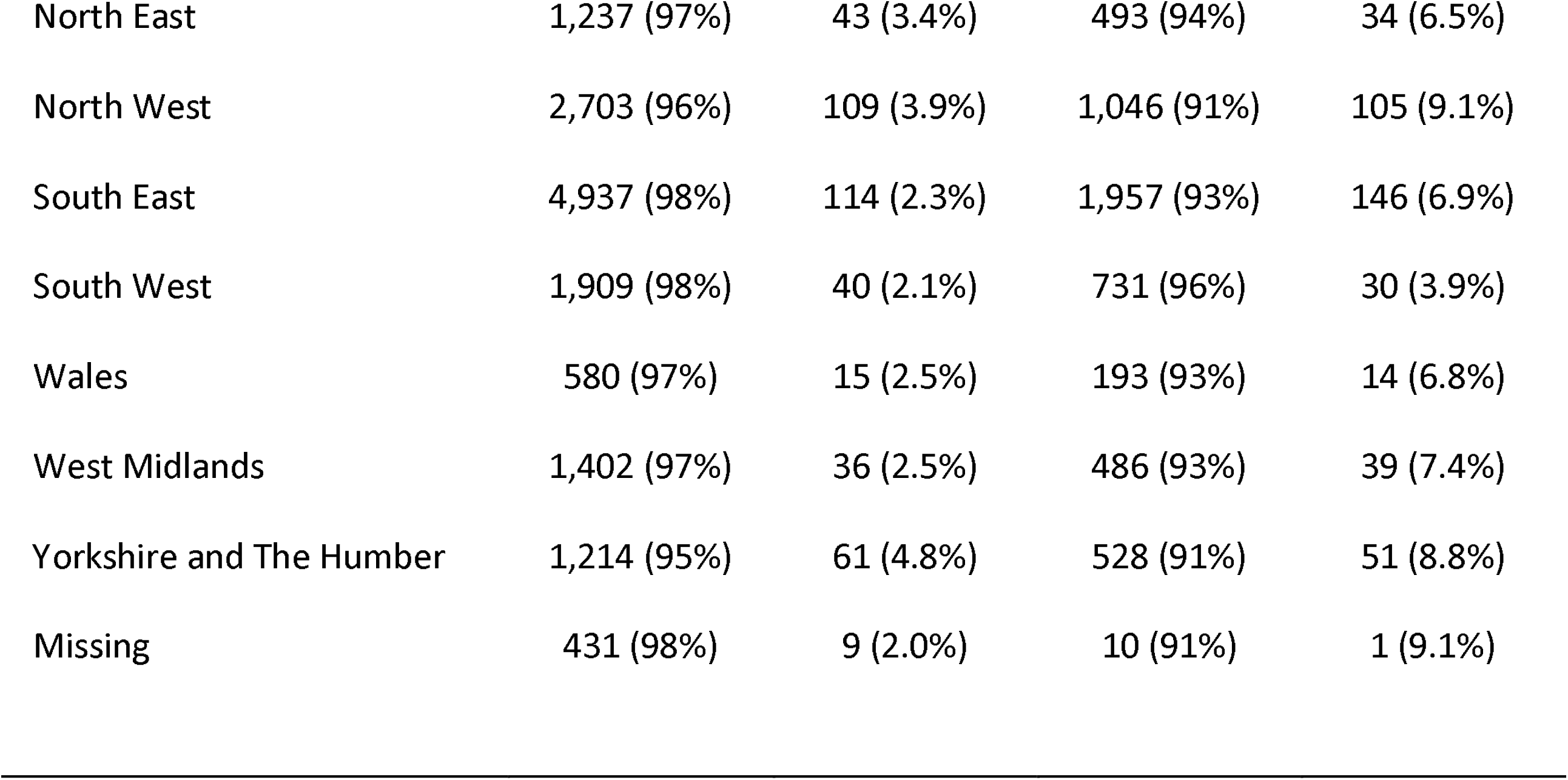
Description of housing characteristics for Virus Watch participants

|  | COVID PCR |  | SARS-CoV2 antibody |  |
| --- | --- | --- | --- | --- |
|  | Negative / Not tested | Positive | Negative | Positive |
| **All** | 25,527 | 867 | 9,483 | 847 |
| **Household overcrowding category** |  |  |  |  |
| under occupied | 22,537 (97%) | 682 (2.9%) | 8,872 (92%) | 733 (7.6%) |
| balanced | 1,961 (95%) | 112 (5.4%) | 454 (86%) | 73 (14%) |
| overcrowded | 1,029 (93%) | 73 (6.6%) | 157 (79%) | 41 (21%) |
| **Age Group** |  |  |  |  |
| 0-15 | 2,288 (97%) | 68 (2.9%) | - | - |
| 16-24 | 1,058 (96%) | 47 (4.3%) | 132 (85%) | 24 (15%) |
| 25-44 | 3,336 (95%) | 180 (5.1%) | 951 (86%) | 155 (14%) |
| 45-64 | 9,013 (96%) | 363 (3.9%) | 3,650 (90%) | 403 (9.9%) |
| 65+ | 9,832 (98%) | 209 (2.1%) | 4,750 (95%) | 265 (5.3%) |
| **Sex** |  |  |  |  |
| Male | 11,385 (97%) | 362 (3.1%) | 4,089 (92%) | 349 (7.9%) |
| Female | 13,656 (96%) | 497 (3.5%) | 5,380 (92%) | 496 (8.4%) |
| Missing | 486 (98%) | 8 (1.6%) | 14 (88%) | 2 (12%) |
| **Ethnicity** |  |  |  |  |
| White British | 22,021 (97%) | 714 (3.1%) | 8,737 (92%) | 728 (7.7%) |
| White Irish | 357 (96%) | 13 (3.5%) | 97 (89%) | 12 (11%) |
| White Other | 1,200 (96%) | 47 (3.8%) | 374 (88%) | 51 (12%) |
| Mixed | 399 (94%) | 25 (5.9%) | 64 (81%) | 15 (19%) |
| South Asian | 562 (94%) | 37 (6.2%) | 100 (78%) | 29 (22%) |
| Other Asian | 180 (94%) | 11 (5.8%) | 55 (92%) | 5 (8.3%) |
| Black | 108 (96%) | 5 (4.4%) | 19 (90%) | 2 (9.5%) |
| Other Ethnicity | 100 (94%) | 6 (5.7%) | 28 (90%) | 3 (9.7%) |
| Prefer not to say | 45 (96%) | 2 (4.3%) | 9 (82%) | 2 (18%) |
| Missing | 555 (99%) | 7 (1.2%) | - | - |
| **Household income combined for last 12 months** |  |  |  |  |
| £0-9,999 | 859 (96%) | 32 (3.6%) | 355 (91%) | 33 (8.5%) |
| £10,000-24,999 | 4,568 (96%) | 174 (3.7%) | 2,225 (94%) | 149 (6.3%) |
| £25,000-49,999 | 7,367 (97%) | 245 (3.2%) | 3,421 (92%) | 290 (7.8%) |
| £50,000-£74,999 | 4,036 (96%) | 163 (3.9%) | 1,547 (90%) | 173 (10%) |
| £75,000-£99,999 | 2,052 (97%) | 71 (3.3%) | 690 (90%) | 79 (10%) |
| £100,000+ | 2,155 (97%) | 71 (3.2%) | 640 (88%) | 87 (12%) |
| Prefer not to say | 1,350 (97%) | 38 (2.7%) | 605 (94%) | 36 (5.6%) |
| Missing | 3,140 (98%) | 73 (2.3%) | - | - |
| **Geographical region** |  |  |  |  |
| East Midlands | 2,268 (96%) | 84 (3.6%) | 822 (92%) | 68 (7.6%) |
| East of England | 5,624 (97%) | 184 (3.2%) | 2,318 (94%) | 160 (6.5%) |
| London | 3,222 (95%) | 172 (5.1%) | 899 (82%) | 199 (18%) |
| North East | 1,237 (97%) | 43 (3.4%) | 493 (94%) | 34 (6.5%) |
| North West | 2,703 (96%) | 109 (3.9%) | 1,046 (91%) | 105 (9.1%) |
| South East | 4,937 (98%) | 114 (2.3%) | 1,957 (93%) | 146 (6.9%) |
| South West | 1,909 (98%) | 40 (2.1%) | 731 (96%) | 30 (3.9%) |
| Wales | 580 (97%) | 15 (2.5%) | 193 (93%) | 14 (6.8%) |
| West Midlands | 1,402 (97%) | 36 (2.5%) | 486 (93%) | 39 (7.4%) |
| Yorkshire and The Humber | 1,214 (95%) | 61 (4.8%) | 528 (91%) | 51 (8.8%) |
| Missing | 431 (98%) | 9 (2.0%) | 10 (91%) | 1 (9.1%) |

**Table 3.**
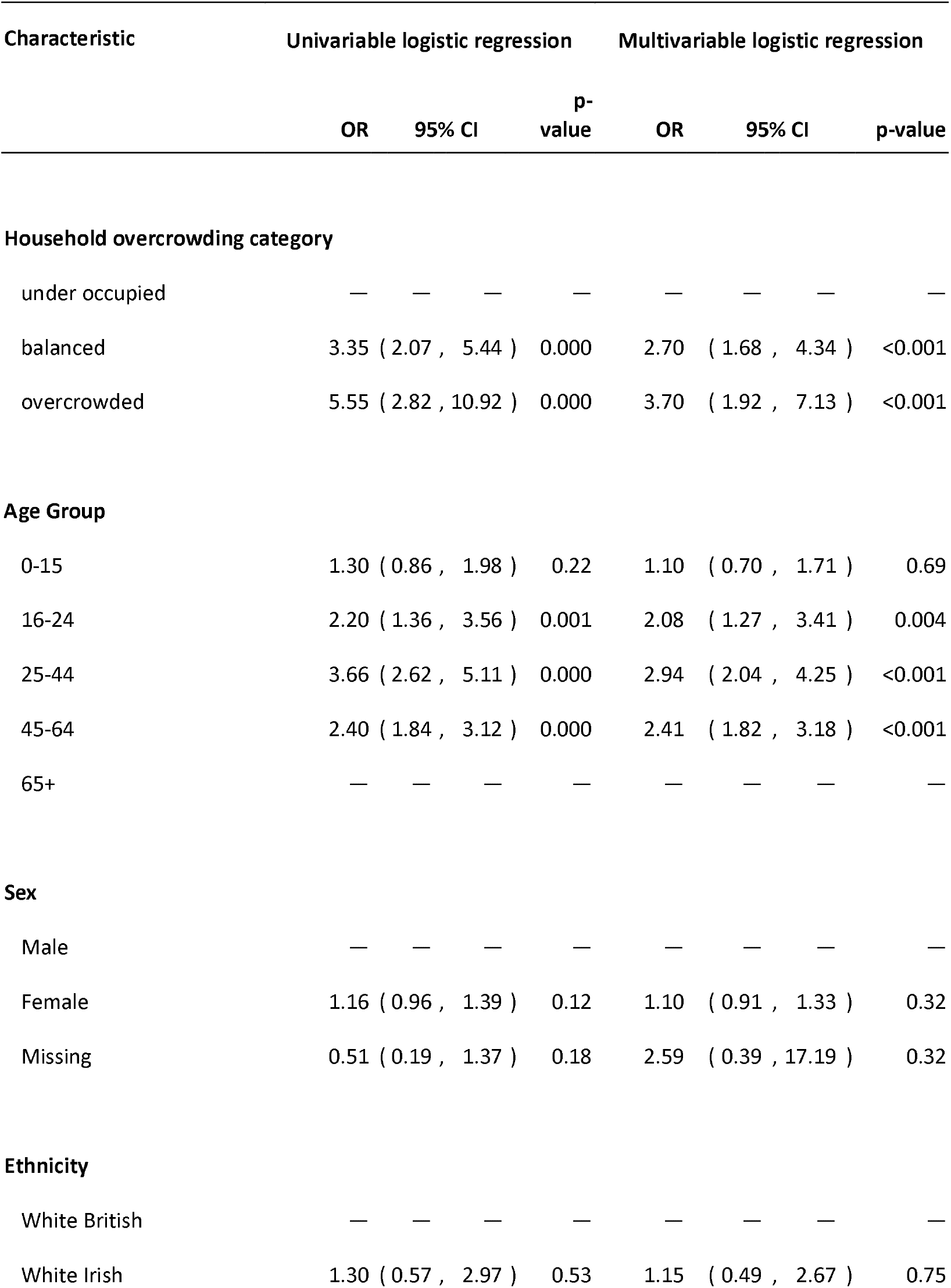
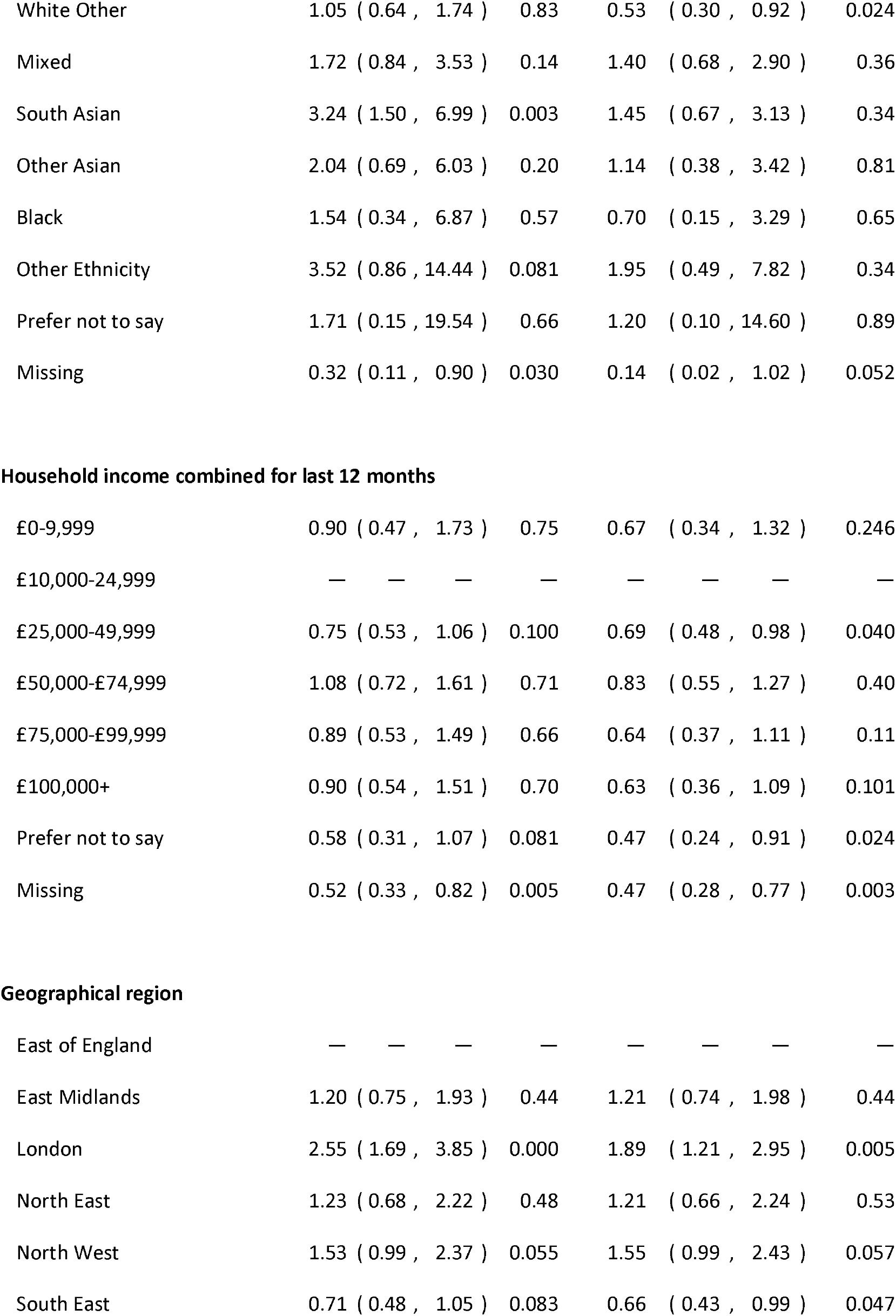
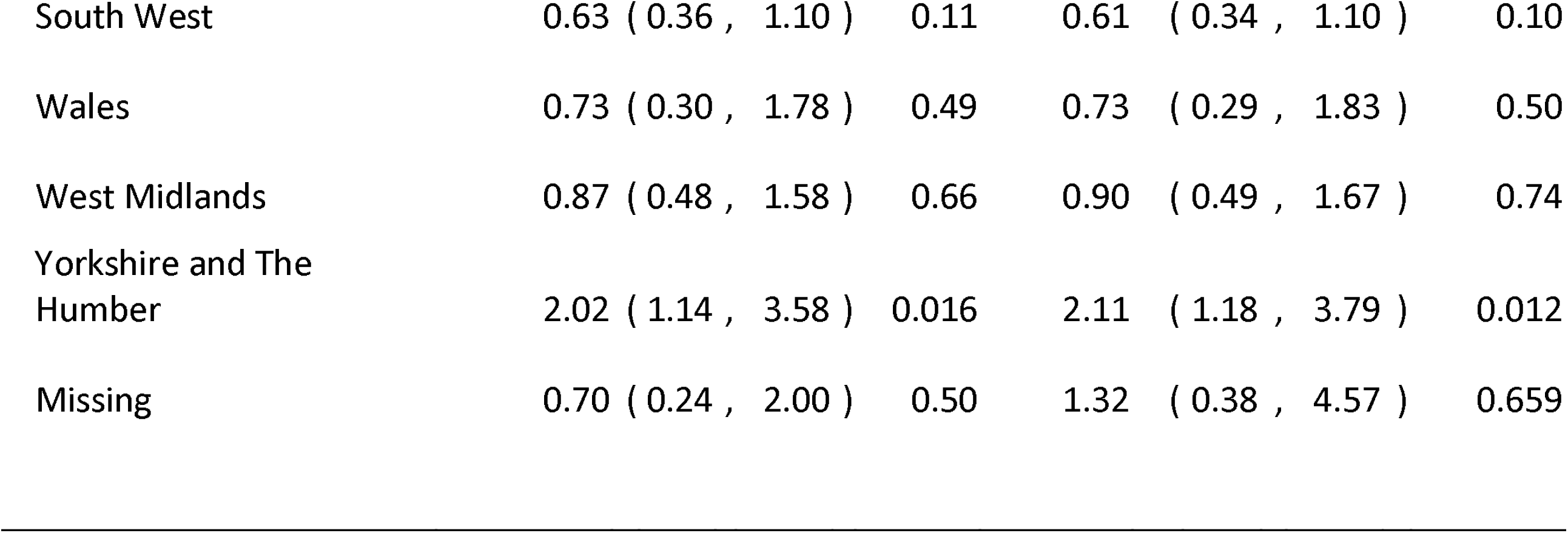
Univariable and multivariable mixed effects logistic regression models examining the association between household overcrowding and SARS-CoV-2 PCR test results.

| Characteristic | Univariable logistic regression |  |  |  | Multivariable logistic regression |  |  |  |
| --- | --- | --- | --- | --- | --- | --- | --- | --- |
|  | OR | 95% CI |  | p-value | OR | 95% CI |  | p-value |
| **Household overcrowding category** |  |  |  |  |  |  |  |  |
| under occupied | — | — | — | — | — | — | — | — |
| balanced | 3.35 | ( 2.07 , 5.44 ) |  | 0.000 | 2.70 | ( 1.68 , 4.34 ) |  | <0.001 |
| overcrowded | 5.55 | ( 2.82 , 10.92 ) |  | 0.000 | 3.70 | ( 1.92 , 7.13 ) |  | <0.001 |
| **Age Group** |  |  |  |  |  |  |  |  |
| 0-15 | 1.30 | ( 0.86 , 1.98 ) |  | 0.22 | 1.10 | ( 0.70 , 1.71 ) |  | 0.69 |
| 16-24 | 2.20 | ( 1.36 , 3.56 ) |  | 0.001 | 2.08 | ( 1.27 , 3.41 ) |  | 0.004 |
| 25-44 | 3.66 | ( 2.62 , 5.11 ) |  | 0.000 | 2.94 | ( 2.04 , 4.25 ) |  | <0.001 |
| 45-64 | 2.40 | ( 1.84 , 3.12 ) |  | 0.000 | 2.41 | ( 1.82 , 3.18 ) |  | <0.001 |
| 65+ | — | — | — | — | — | — | — | — |
| **Sex** |  |  |  |  |  |  |  |  |
| Male | — | — | — | — | — | — | — | — |
| Female | 1.16 | ( 0.96 , 1.39 ) |  | 0.12 | 1.10 | ( 0.91 , 1.33 ) |  | 0.32 |
| Missing | 0.51 | ( 0.19 , 1.37 ) |  | 0.18 | 2.59 | ( 0.39 , 17.19 ) |  | 0.32 |
| **Ethnicity** |  |  |  |  |  |  |  |  |
| White British | — | — | — | — | — | — | — | — |
| White Irish | 1.30 | ( 0.57 , 2.97 ) |  | 0.53 | 1.15 | ( 0.49 , 2.67 ) |  | 0.75 |

|  |  |  |  |  |
| --- | --- | --- | --- | --- |
| White Other | 1.05 ( 0.64 , 1.74 ) | 0.83 | 0.53 ( 0.30 , 0.92 ) | 0.024 |
| Mixed | 1.72 ( 0.84 , 3.53 ) | 0.14 | 1.40 ( 0.68 , 2.90 ) | 0.36 |
| South Asian | 3.24 ( 1.50 , 6.99 ) | 0.003 | 1.45 ( 0.67 , 3.13 ) | 0.34 |
| Other Asian | 2.04 ( 0.69 , 6.03 ) | 0.20 | 1.14 ( 0.38 , 3.42 ) | 0.81 |
| Black | 1.54 ( 0.34 , 6.87 ) | 0.57 | 0.70 ( 0.15 , 3.29 ) | 0.65 |
| Other Ethnicity | 3.52 ( 0.86 , 14.44 ) | 0.081 | 1.95 ( 0.49 , 7.82 ) | 0.34 |
| Prefer not to say | 1.71 ( 0.15 , 19.54 ) | 0.66 | 1.20 ( 0.10 , 14.60 ) | 0.89 |
| Missing | 0.32 ( 0.11 , 0.90 ) | 0.030 | 0.14 ( 0.02 , 1.02 ) | 0.052 |

Household income combined for last 12 months
|  |  |  |  |  |
| --- | --- | --- | --- | --- |
| £0-9,999 | 0.90 ( 0.47 , 1.73 ) | 0.75 | 0.67 ( 0.34 , 1.32 ) | 0.246 |
| £10,000-24,999 | — — — — | — | — — — | — |
| £25,000-49,999 | 0.75 ( 0.53 , 1.06 ) | 0.100 | 0.69 ( 0.48 , 0.98 ) | 0.040 |
| £50,000-£74,999 | 1.08 ( 0.72 , 1.61 ) | 0.71 | 0.83 ( 0.55 , 1.27 ) | 0.40 |
| £75,000-£99,999 | 0.89 ( 0.53 , 1.49 ) | 0.66 | 0.64 ( 0.37 , 1.11 ) | 0.11 |
| £100,000+ | 0.90 ( 0.54 , 1.51 ) | 0.70 | 0.63 ( 0.36 , 1.09 ) | 0.101 |
| Prefer not to say | 0.58 ( 0.31 , 1.07 ) | 0.081 | 0.47 ( 0.24 , 0.91 ) | 0.024 |
| Missing | 0.52 ( 0.33 , 0.82 ) | 0.005 | 0.47 ( 0.28 , 0.77 ) | 0.003 |

Geographical region
|  |  |  |  |  |
| --- | --- | --- | --- | --- |
| East of England | — — — — | — | — — — | — |
| East Midlands | 1.20 ( 0.75 , 1.93 ) | 0.44 | 1.21 ( 0.74 , 1.98 ) | 0.44 |
| London | 2.55 ( 1.69 , 3.85 ) | 0.000 | 1.89 ( 1.21 , 2.95 ) | 0.005 |
| North East | 1.23 ( 0.68 , 2.22 ) | 0.48 | 1.21 ( 0.66 , 2.24 ) | 0.53 |
| North West | 1.53 ( 0.99 , 2.37 ) | 0.055 | 1.55 ( 0.99 , 2.43 ) | 0.057 |
| South East | 0.71 ( 0.48 , 1.05 ) | 0.083 | 0.66 ( 0.43 , 0.99 ) | 0.047 |
| South West | 0.63 ( 0.36 , 1.10 ) | 0.11 | 0.61 ( 0.34 , 1.10 ) | 0.10 |
| Wales | 0.73 ( 0.30 , 1.78 ) | 0.49 | 0.73 ( 0.29 , 1.83 ) | 0.50 |
| West Midlands | 0.87 ( 0.48 , 1.58 ) | 0.66 | 0.90 ( 0.49 , 1.67 ) | 0.74 |
| Yorkshire and The<br>Humber | 2.02 ( 1.14 , 3.58 ) | 0.016 | 2.11 ( 1.18 , 3.79 ) | 0.012 |
| Missing | 0.70 ( 0.24 , 2.00 ) | 0.50 | 1.32 ( 0.38 , 4.57 ) | 0.659 |

As of 3rd May 2021 we had SARS-CoV-2 anti-N total immunoglobulin assay antibody results for 10,330 participants in the laboratory cohort with housing data (Table 2). The proportion of participants with a positive SARS-CoV-2 antibody result was highest in the overcrowded group (20.7%; 41/198) and lowest in the under-occupied group (7.6%; 733/9,605; Table 2). In a mixed effects logistic regression model that included age, sex, ethnicity, household income and geographical region as fixed effects and a household-level random effect, we found evidence of an increased odds of having a positive SARS-CoV-2 an result in individuals living in overcrowded houses (OR: 2.96; 95% CI: 1.13, 7.74; p-value =0.027) and some evidence of this amongst those living in balanced houses (OR: 1.70; 95% CI: 0.89, 3.25; p-value = 0.11) compared to people living in under-occupied houses (Table 4). The proportion of variation at the household level was 74.0%.

**Table 4.**
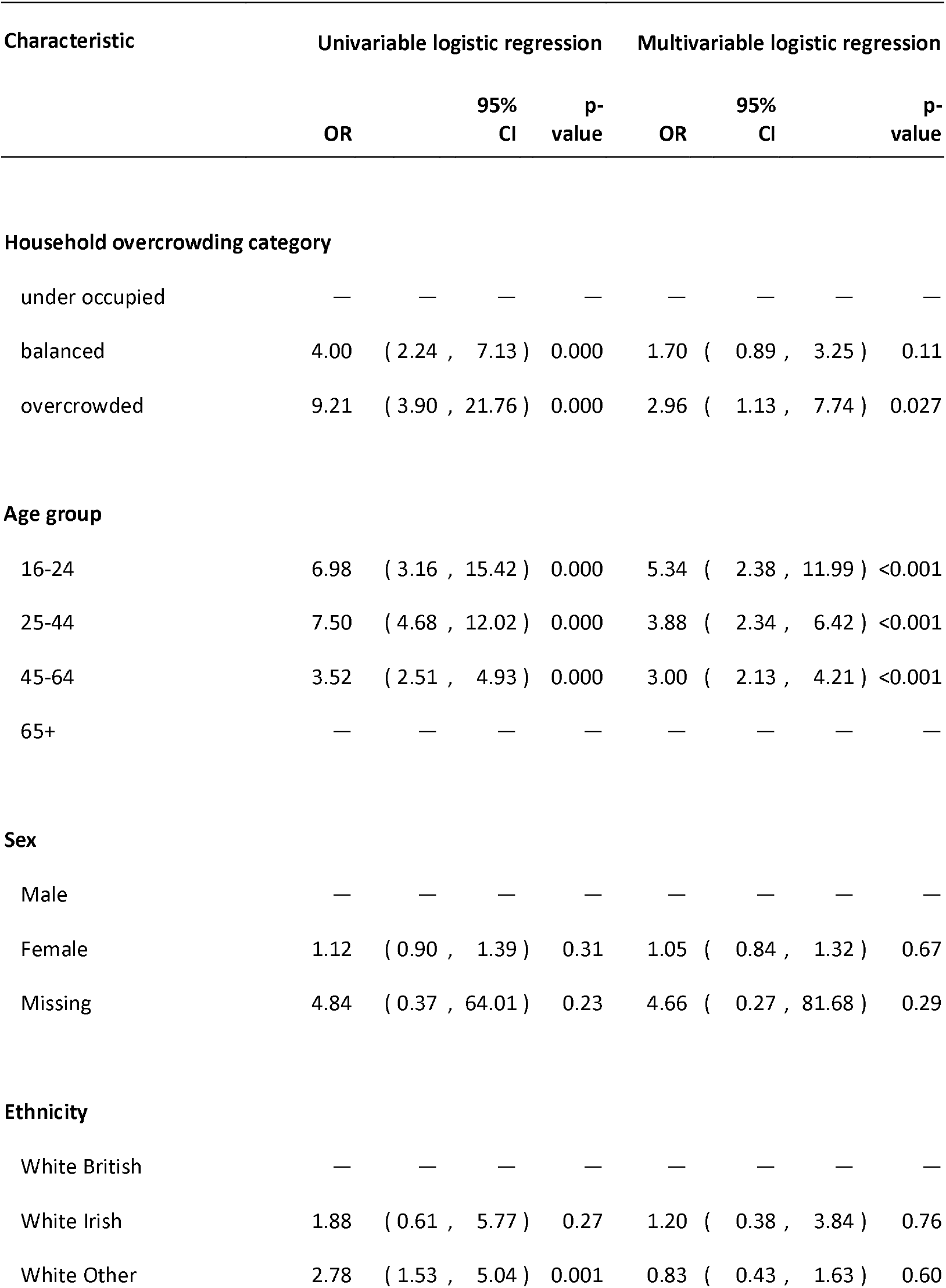
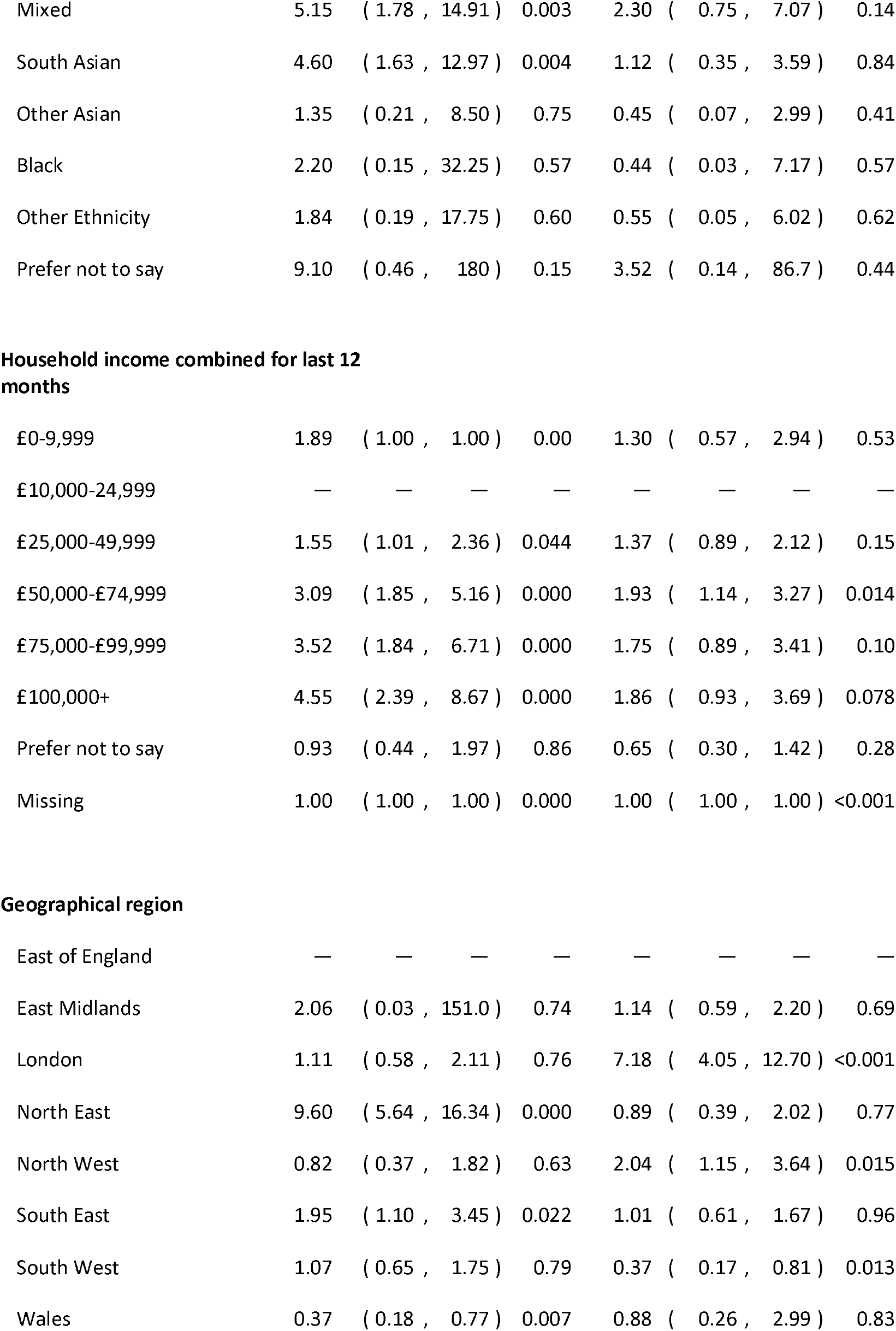
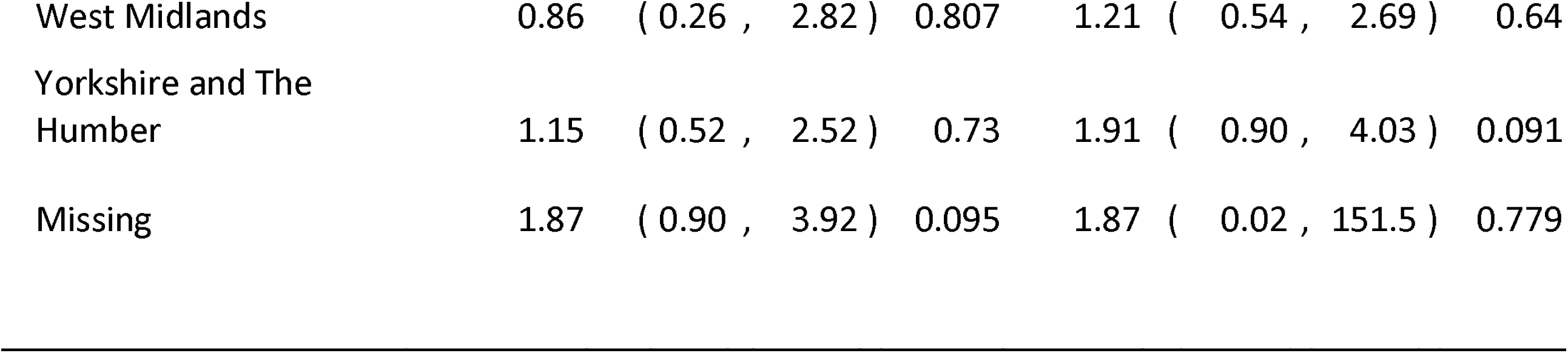
Univariable and multivariable mixed effects logistic regression models examining the association between household overcrowding and SARS-CoV-2 antibody test results.

| Characteristic | Univariable logistic regression |  |  | Multivariable logistic regression |  |  |
| --- | --- | --- | --- | --- | --- | --- |
|  | OR | 95% CI | p-value | OR | 95% CI | p-value |
| **Household overcrowding category** |  |  |  |  |  |  |
| under occupied | — | — | — | — | — | — |
| balanced | 4.00 | ( 2.24 , 7.13 ) | 0.000 | 1.70 | ( 0.89 , 3.25 ) | 0.11 |
| overcrowded | 9.21 | ( 3.90 , 21.76 ) | 0.000 | 2.96 | ( 1.13 , 7.74 ) | 0.027 |
| **Age group** |  |  |  |  |  |  |
| 16-24 | 6.98 | ( 3.16 , 15.42 ) | 0.000 | 5.34 | ( 2.38 , 11.99 ) | <0.001 |
| 25-44 | 7.50 | ( 4.68 , 12.02 ) | 0.000 | 3.88 | ( 2.34 , 6.42 ) | <0.001 |
| 45-64 | 3.52 | ( 2.51 , 4.93 ) | 0.000 | 3.00 | ( 2.13 , 4.21 ) | <0.001 |
| 65+ | — | — | — | — | — | — |
| **Sex** |  |  |  |  |  |  |
| Male | — | — | — | — | — | — |
| Female | 1.12 | ( 0.90 , 1.39 ) | 0.31 | 1.05 | ( 0.84 , 1.32 ) | 0.67 |
| Missing | 4.84 | ( 0.37 , 64.01 ) | 0.23 | 4.66 | ( 0.27 , 81.68 ) | 0.29 |
| **Ethnicity** |  |  |  |  |  |  |
| White British | — | — | — | — | — | — |
| White Irish | 1.88 | ( 0.61 , 5.77 ) | 0.27 | 1.20 | ( 0.38 , 3.84 ) | 0.76 |
| White Other | 2.78 | ( 1.53 , 5.04 ) | 0.001 | 0.83 | ( 0.43 , 1.63 ) | 0.60 |
| Mixed | 5.15 | ( 1.78 , 14.91 ) | 0.003 | 2.30 | ( 0.75 , 7.07 ) | 0.14 |
| South Asian | 4.60 | ( 1.63 , 12.97 ) | 0.004 | 1.12 | ( 0.35 , 3.59 ) | 0.84 |
| Other Asian | 1.35 | ( 0.21 , 8.50 ) | 0.75 | 0.45 | ( 0.07 , 2.99 ) | 0.41 |
| Black | 2.20 | ( 0.15 , 32.25 ) | 0.57 | 0.44 | ( 0.03 , 7.17 ) | 0.57 |
| Other Ethnicity | 1.84 | ( 0.19 , 17.75 ) | 0.60 | 0.55 | ( 0.05 , 6.02 ) | 0.62 |
| Prefer not to say | 9.10 | ( 0.46 , 180 ) | 0.15 | 3.52 | ( 0.14 , 86.7 ) | 0.44 |
| Household income combined for last 12 months |  |  |  |  |  |  |
| £0-9,999 | 1.89 | ( 1.00 , 1.00 ) | 0.00 | 1.30 | ( 0.57 , 2.94 ) | 0.53 |
| £10,000-24,999 | — | — | — | — | — | — |
| £25,000-49,999 | 1.55 | ( 1.01 , 2.36 ) | 0.044 | 1.37 | ( 0.89 , 2.12 ) | 0.15 |
| £50,000-£74,999 | 3.09 | ( 1.85 , 5.16 ) | 0.000 | 1.93 | ( 1.14 , 3.27 ) | 0.014 |
| £75,000-£99,999 | 3.52 | ( 1.84 , 6.71 ) | 0.000 | 1.75 | ( 0.89 , 3.41 ) | 0.10 |
| £100,000+ | 4.55 | ( 2.39 , 8.67 ) | 0.000 | 1.86 | ( 0.93 , 3.69 ) | 0.078 |
| Prefer not to say | 0.93 | ( 0.44 , 1.97 ) | 0.86 | 0.65 | ( 0.30 , 1.42 ) | 0.28 |
| Missing | 1.00 | ( 1.00 , 1.00 ) | 0.000 | 1.00 | ( 1.00 , 1.00 ) | <0.001 |
| Geographical region |  |  |  |  |  |  |
| East of England | — | — | — | — | — | — |
| East Midlands | 2.06 | ( 0.03 , 151.0 ) | 0.74 | 1.14 | ( 0.59 , 2.20 ) | 0.69 |
| London | 1.11 | ( 0.58 , 2.11 ) | 0.76 | 7.18 | ( 4.05 , 12.70 ) | <0.001 |
| North East | 9.60 | ( 5.64 , 16.34 ) | 0.000 | 0.89 | ( 0.39 , 2.02 ) | 0.77 |
| North West | 0.82 | ( 0.37 , 1.82 ) | 0.63 | 2.04 | ( 1.15 , 3.64 ) | 0.015 |
| South East | 1.95 | ( 1.10 , 3.45 ) | 0.022 | 1.01 | ( 0.61 , 1.67 ) | 0.96 |
| South West | 1.07 | ( 0.65 , 1.75 ) | 0.79 | 0.37 | ( 0.17 , 0.81 ) | 0.013 |
| Wales | 0.37 | ( 0.18 , 0.77 ) | 0.007 | 0.88 | ( 0.26 , 2.99 ) | 0.83 |
| West Midlands | 0.86 | ( 0.26 , 2.82 ) | 0.807 | 1.21 | ( 0.54 , 2.69 ) | 0.64 |
| Yorkshire and The<br>Humber | 1.15 | ( 0.52 , 2.52 ) | 0.73 | 1.91 | ( 0.90 , 4.03 ) | 0.091 |
| Missing | 1.87 | ( 0.90 , 3.92 ) | 0.095 | 1.87 | ( 0.02 , 151.5 ) | 0.779 |

Sensitivity analyses were consistent with the finding that overcrowding was associated with increased risk of PCR and anti-N total immunoglobulin assay antibody confirmed SARS-CoV-2 (Table 5). In a sensitivity analysis where we did not subtract 1 from the number of rooms there was evidence of an increased odds of having a positive SARS-CoV-2 antigen result in balanced (OR: 3.37; 95% CI: 1.52, 7.45; p-value = 0.003) and overcrowded households (OR: 2.53; 95% CI: 0.96, 6.62; p-value = 0.059) compared to those in under-occupied houses.

**Table 5.**
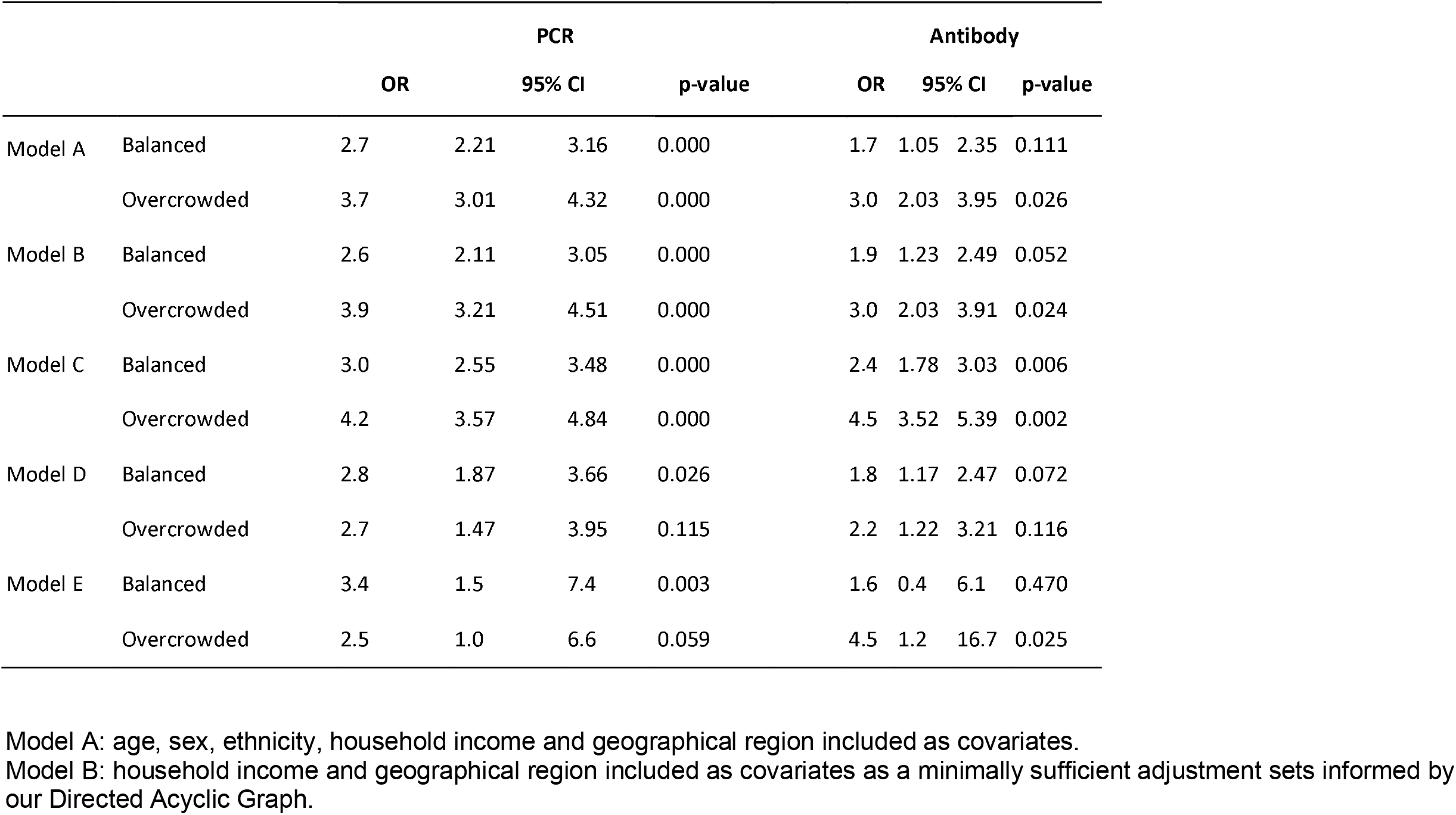
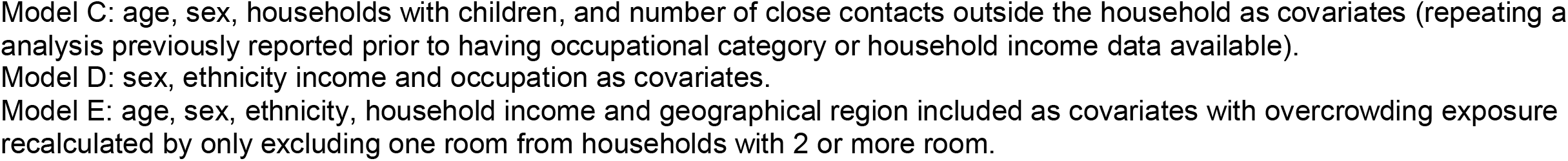
Sensitivity analyses of mixed effects logistic regression models.

## Discussion

We estimate that overcrowded households have between two to four times the odds of PCR confirmed SARS-CoV-2 and anti-N total immunoglobulin assay antibody positivity compared to under-occupied households. This elevated odds of confirmed SARS-CoV-2 remained after we adjusted for differences in demographic and other socio-economic factors between households that may increase risk of being infected. We also found that people in accommodation considered balanced - where the number of rooms was equal to the number of people - were at increased risk of PCR confirmed SARS-CoV-2 compared to under-occupied houses.

Virus Watch is a large national household community cohort study of the occurrence and risk factors for SARS-CoV-2 infection and is designed to estimate incidence of PCR confirmed SARS-CoV-2 and measure effectiveness and impact of recommended COVID-19 control measures. Individuals in the study are geographically distributed across England and Wales and the cohort is diverse in terms of age, sex, ethnicity and socio-economic composition, and levels of overcrowding were comparable with national estimates for England and Wales.

There were more participants taking part in the February 2021 monthly survey that were over the age of 65 compared to the general population in England and Wales, more people of White British ethnicity, and more people of household size 2. Virus Watch is limited by the fact that only households with a lead householder able to speak English and access the internet were able to take part in the study.^16^ An important additional limitation is that only households of up to six people were eligible for inclusion. By not including households of over six people we are likely underestimating the risk associated with overcrowding. Conversely, we cannot rule out residual confounding as a possible alternative explanation for some of the excess risk in the associations we describe.

Access to SARS-CoV-2 antigen testing is socio-economically patterned, with those in more deprived areas having less ability to access tests and less likely to be contact traced.^23^ Our data may therefore under-estimate the true infection risk associated with overcrowded housing. To mitigate this bias in access to antigen testing, we analysed antibody data from the laboratory cohort where home test kits were provided to all adult participants.

Our findings highlight the importance of public health interventions to prevent and stop the spread of SARS-CoV-2 considering the much greater risk of being infected for people living in overcrowded households. The pathways between overcrowding and increased risk of infection are not fully understood.^10^ Overcrowded households are more likely to be found in socioeconomically deprived and urban areas, and higher levels of COVID-19 in these communities include higher likelihood of working in public-facing and essential occupations^24^, lower likelihood of working from home during lockdowns, constraints to self-isolation when ill or in contact with a case (both financial and due to lack of suitable space within an overcrowded house), increased number of household members sharing of common spaces and facilities. Overcrowded households are also more to suffer from environmental exposures that have been linked to COVID-19 including air pollution.^12^

Overcrowding is an important risk through which COVID-19 inequalities manifest. To address social, ethnic and regional inequalities in COVID-19 outcomes, public health responses should explicitly address overcrowded housing. Infectious cases in overcrowded houses are less likely to be able to isolate from other household members or avoid using shared spaces including bathrooms. However, existing advice on the importance of infectious cases using masks and ventilation through opening of windows should be supported by tailored communications strategies.^25^ Schemes that provide hotel accommodation for cases with vulnerable residents in overcrowded households, such as those used internationally^26–28^ and recently introduced in the London Borough of Newham^29^, should be considered a priority public health intervention. Such community quarantine and isolation options require clear, inclusive guidance with specific advice for large households and multigenerational families.^10^ Although the UK does not target COVID-19 vaccine according to social factors, our results highlight the need to ensure high vaccine coverage in areas with high levels of overcrowding. There is currently evidence of lower COVID-19 vaccination rates in such areas,^30^ highlighting the importance of working closely with local community groups to increase vaccination intention and ensuring vaccination is highly accessible.

Improving the standard and supply of housing in the UK is important for the COVID-19 recovery as 32% of all households are estimated to experience overcrowding, affordability challenges or non-decent housing that influence their risk.^31^ In relation to these challenges, a paper by the ethnicity sub-group of the Scientific Advisory Group for Emergencies (SAGE) recommends the following measures to reduce the risk of transmission within households: provision of emergency grants for repair and maintenance of social and private rental housing, particularly to increase ventilation; removing the benefit cap and welfare restrictions, particularly in high housing cost areas and for immigrant families; reviewing implementation of the bedroom tax for those in multi-person households; and investment in affordable childcare and alternative community spaces for social connection, particularly for the elderly.^10^

Addressing England’s overcrowding challenge is complex and requires action from multiple government and private sector stakeholders. A recent House of Commons Library briefing report has highlighted actions related to the following: revising the statutory housing standard, increasing the housing supply and size of homes/rooms, and measures to reduce under-occupation in social housing.^32^ Breaching the statutory overcrowding standard is a criminal offence, yet the threshold is considered to be too low such that very few homes are statutorily overcrowded, reducing local authorities’ ability to take action on this problem.^32^ Another significant challenge is that housing supply has fallen short of demand for decades.^33^ There are a wide range of factors affecting housing supply (and therefore also influencing affordability and overcrowding in England) including: construction capacity, planning regulation, public opposition to new development, overseas buyers, overreliance on the private sector and underuse of social approaches to housing development, among other factors.^34^ The size of English homes may also be an important factor as they are among the smallest in Europe^35^ and the Nationally Described Space Standard^36^ for new homes is only mandatory if implemented through local planning policies following tests for need and viability. Finally, although there are under-occupied and empty homes in England, these are not suitable to solve the housing supply problem due to their location, ownership and other factors.^37^

Housing is an important determinant of health for many physical and mental health conditions and COVID-19 has exposed, in real-time, vulnerabilities in the English housing stock, particularly for low-income and minority ethnic populations. Looking forward, the country’s health and resilience to future pandemics and other risks, such as climate change^38,39^, are dependent on the state of the housing stock. This analysis of overcrowding and COVID-19 underscores the need to improve housing for health. As part of the agenda to Build Back Better^40^ and fairer^41–43^, investment in sustainable, high-quality and more affordable housing will support health, jobs and the wider economic recovery so that we are prepared for the key challenges of the 21st century.

## Data Availability

We aim to share aggregate data from this project on our website and via a “Findings so far” section on our website - https://ucl-virus-watch.net/. We will also be sharing individual record level data on a research data sharing service such as the Office of National Statistics Secure Research Service. In sharing the data we will work within the principles set out in the UKRI Guidance on best practice in the management of research data. Access to use of the data whilst research is being conducted will be managed by the Chief Investigators (ACH and RWA) in accordance with the principles set out in the UKRI guidance on best practice in the management of research data. We will put analysis code on publicly available repositories to enable their reuse.

## Declarations of interest

ACH serves on the UK New and Emerging Respiratory Virus Threats Advisory Group. AMJ was a Governor of Wellcome Trust from 2011-18 and is Chair of the Committee for Strategic Coordination for Health of the Public Research. SVK is a member of the UK Government’s Scientific Advisory Group on Emergencies (SAGE) subgroup on ethnicity and co-chair of the Scottish Government’s Expert Reference Group on ethnicity and COVID-19.

## Funding

The research costs for the study have been supported by the MRC Grant Ref: MC_PC 19070 awarded to UCL on 30 March 2020 and MRC Grant Ref: MR/V028375/1 awarded on 17 August 2020. The study also received $15,000 of Facebook advertising credit to support a pilot social media recruitment campaign on 18th August 2020. This study was supported by the Wellcome Trust through a Wellcome Clinical Research Career Development Fellowship to RA [206602]. SVK acknowledges funding from a NRS Senior Clinical Fellowship (SCAF/15/02), the Medical Research Council (MC_UU_00022/2) and the Scottish Government Chief Scientist Office (SPHSU17).

## Supplementary Appendices

**Figure S1.**
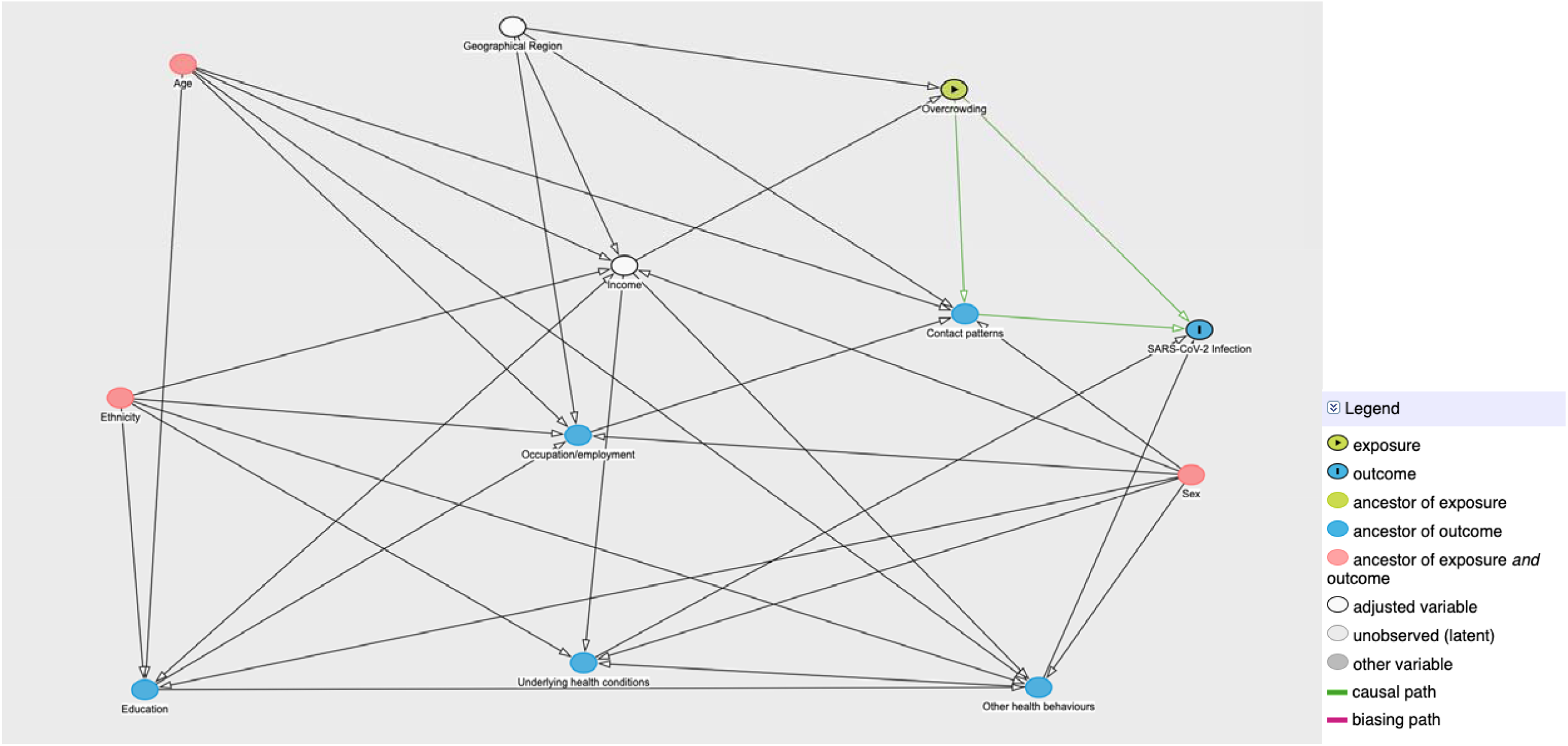
Directed acyclic graph for estimating the total effect of Overcrowding on SARS-CoV-2 Infection.

Housing-related survey questions used in the February 2021 monthly survey.

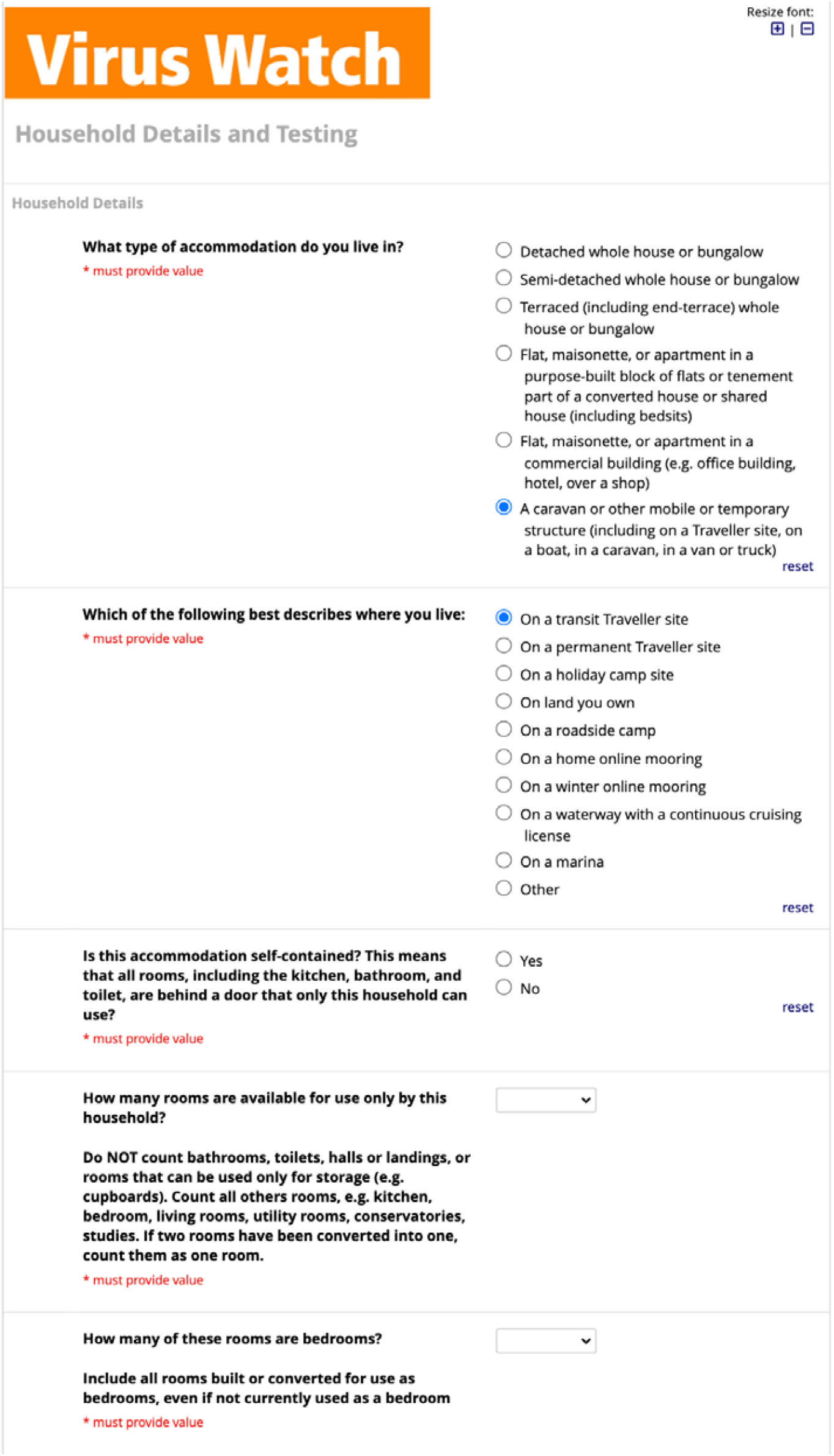
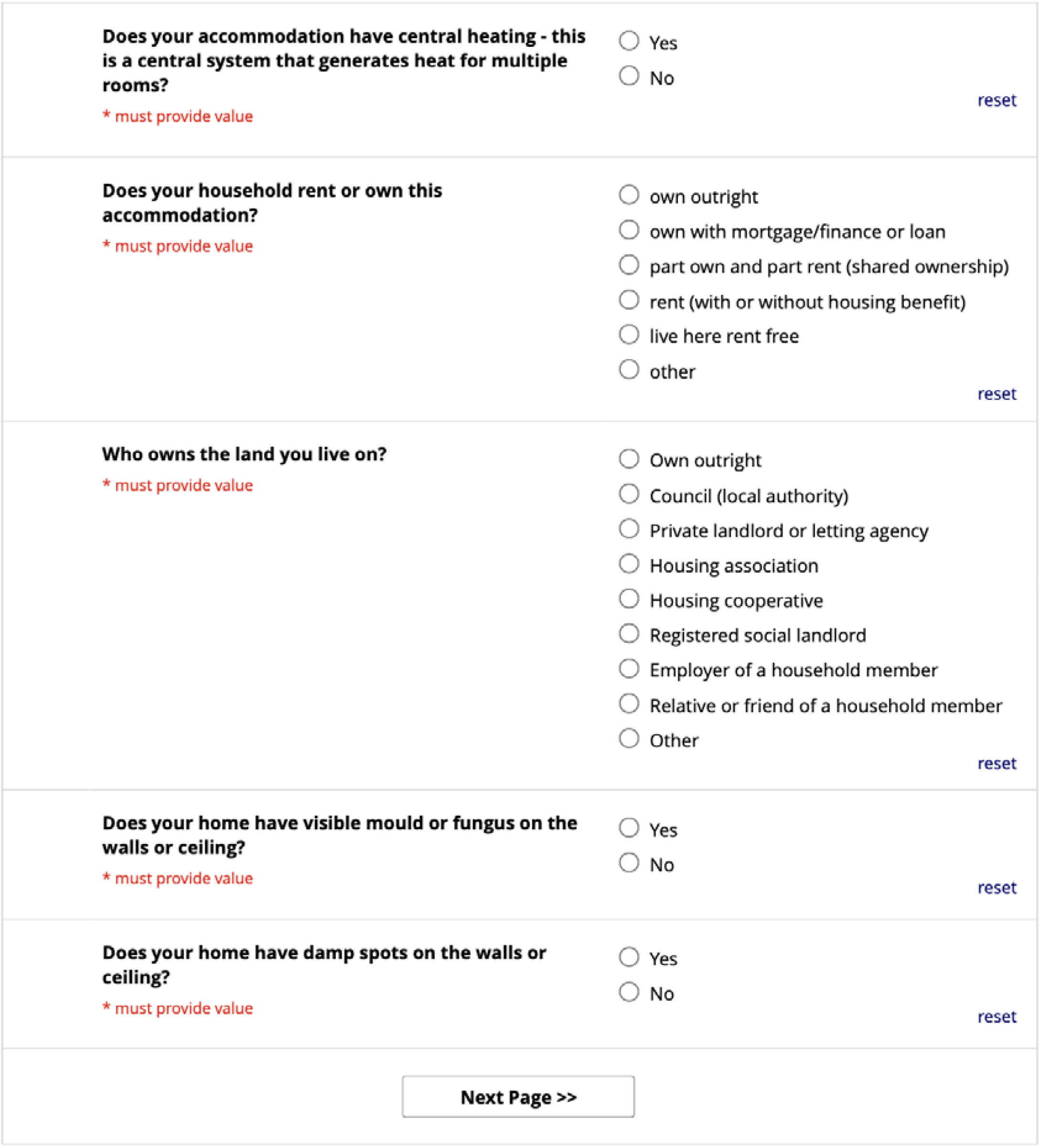

## Notes

### Author Declarations

This study has been approved by the Hampstead NHS Health Research Authority Ethics Committee. Ethics approval number - 20/HRA/2320.

### Summary of Updates

Minor edits to text in the introduction and discussion, including the addition of new literature. Results on the proportion of variation at the household level corrected.

